# Medical Students’ Perceptions of and Attitudes Toward English as a Medium of Instruction at the Faculty of Medicine and Pharmacy of Rabat: A Cross-Sectional Study

**DOI:** 10.64898/2026.01.22.26344560

**Authors:** I Maaroufi, R Razine, Z Zeghari, I Zeddari, D Laayadi, M Obtel, J Belayachi, R Abouqal

**Author notes:** Corresponding author: Maaroufi Ilham.

## Abstract

**Background:** The global adoption of English-medium instruction (EMI) in higher education has introduced complex implementation challenges, whose severity often depends on the resources available within specific educational contexts. Evidence remains limited in public medical schools in under-resourced, non-Anglophone countries, serving a socioeconomically and educationally diverse student population. In such settings, the direct transfer of existing EMI integration models presents significant practical challenges.

**Objective:** This exploratory, observational, monocentric study aimed to investigate medical students’ perceptions and attitudes towards EMI implementation at the Faculty of Medicine and Pharmacy of Rabat (FMPR) in Morocco, to evaluate their readiness for EMI adoption.

**Methods:** A cross-sectional survey was administered to 102 second-curriculum-level medical students at FMPR. The 23-item questionnaire included Likert-type scales and multiple-choice questions across four domains: demographic data, self-reported English language (EL) proficiency, language perceptions and attitudes, and EMI needs. Bivariate, univariate and multivariate logistic regression analyses were conducted to determine the effects of the explanatory variables on EMI choice.

**Results:** The sample comprised 65 females and 37 males. Most participants reported using EL speaking and listening skills “often” on a daily basis, while writing and reading skills were reported as being used “rarely.” Participants rated their general EL proficiency and their academic listening and reading skills as “good.” Overall, 92.2% (94/102) of students were strongly in favor of implementing EMI at FMPR. Bivariate analysis showed significant positive associations between EMI choice and age (*P* = .035), course year (*p* < .001), interest in learning EL (*P* = .011), and the belief that EL should be the language of higher education (*P* < .001). Preference for EMI increased alongside course year, peaking at 96.4% among students in the third and fourth years. Multivariate logistic regression confirmed that being in course years 3 and 4 (OR = 8.83, 95% CI [2.05–172.69], *P* = .009), having an interest in learning EL (OR = 6.90, 95% CI [1.31–36.16], *P* = .022), and believing that EL should be the language of higher education (OR = 6.38, 95% CI [1.87–21.72], *P* =.003) were significant positive predictors of EMI choice.

**Conclusions:** Medical students at FMPR exhibit highly positive perceptions and attitudes toward the potential implementation of EMI, despite variations in their self-reported EL readiness. These findings provide actionable insights for the successful integration of EMI that extend beyond the Moroccan context, offering a valuable framework for medical institutions in other under-resourced, low- and middle-income, non-Anglophone settings across the Global South.

## Introduction

The twenty-first century is witnessing the establishment of English language (EL) as the language of globalization. EL has gained global acceptance across diverse fields, serving as the primary medium for trade, diplomacy, technology, tourism, scientific research, medicine, and education [1]. In higher education, EL has been increasingly adopted as a medium of instruction in non-Anglophone countries. Macaro defined this pedagogical approach as “the use of EL to teach academic subjects (other than English itself) in countries where the first language of the majority of the population is not English” [2]. Globally, the expanding adoption of English as Medium of Instruction (EMI) programs is driven by rationales such as access to global communication, students’ need for high employability in the global market, improved quality of education and research leading to better university rankings, and fostered cultural understanding [3,4]. Previous research, including British Council surveys, indicates that private educational institutions dominate EMI adoption across various income levels [5]. Conversely, public medical institutions in middle-income regions [7–11] face significantly greater implementation challenges compared to high-income countries [12,13,14]. The present study at the Faculty of Medicine and Pharmacy of Rabat (FMPR) addresses this empirical gap by exploring the potential opportunities and barriers for EMI integration within public medical schools in under-resourced, low- and middle-income, non-Anglophone settings [6,9].

This global trend toward EMI adoption extends beyond former British colonies, where EL is widely used, into linguistically diverse non-Anglophone regions. In Africa, even countries formerly colonized by France, Germany, and Portugal have adopted EMI as a gateway to international medical education [4,15,16]. In the MENA region, Middle Eastern oil economies with Arabic as the official language (AL) and large expatriate populations are transitioning from oil-based to knowledge-based economies, implementing EMI in medical schools despite the challenges stemming from AL-based high school education [8,17,18,19]. North African countries, such as Tunisia and Algeria, where the French language (FL) holds a prominent place in education due to colonial legacy, are progressively adopting EMI in medical schools to meet international standards while retaining French as the primary medium [10,20]. Similarly, Libya shows a modest but growing interest in introducing EMI into medical curricula despite AL being the main medium of instruction in higher education, whereas Egypt’s medical schools largely utilize EMI due to historical British educational influence [21].

Within this regional landscape, Morocco is strategically advancing the status of EL within its educational system. The country initiated an educational reform, commonly referred to as the 2022–2026 Roadmap, to enhance the educational system’s competitiveness through the development of student and educator EL skills [22]. Many universities are currently diversifying their pedagogical offerings with EL programs to attract both Moroccan and international students, honoring Morocco’s commitment to internationalization. Despite these regional and institutional advances, research on EMI in Moroccan public medical faculties remains notably scarce. Ben Hammou examined student experiences in a private EMI health sciences program at Mohammed VI University of Health Sciences [7]. Ben Hammou and Razkane explored the transition to EMI in Moroccan higher education institutions through the perspectives of lecturers in business administration and medicine [23].

To the best of our knowledge, no studies have examined students’ perceptions, attitudes, or EL proficiency needs specifically within public faculties of medicine and pharmacy, such as FMPR. Unlike more privileged private educational contexts, FMPR attracts a socioeconomically diverse population, many of whom lack the financial resources required for private EL proficiency improvement. This presents a critical research gap with profound implications for equitable access to globalized medical education.

This study positions FMPR as a critical case study to explore EMI implementation challenges in public medical education. While situated in Morocco, the issues addressed are highly relevant to similar contexts, namely, low- and middle-income postcolonial countries with non-Anglophone legacies, where public faculties lag behind private institutions in EMI adoption. This disparity is increasingly evident in the Global South, particularly as faculties face a growing need to compete internationally. These demands are illustrated by accreditation pressures from the World Federation for Medical Education (WFME), which requires international-standard EL proficiency for global diploma recognition, and the Educational Commission for Foreign Medical Graduates (ECFMG), which mandated WFME accreditation starting in 2024 for Moroccan graduates to access the United States Medical Licensing Examination (USMLE) [24,25].

To explore the potential for EMI integration in Moroccan public medical faculties, this study aims to investigate medical students’ perceptions and attitudes toward EMI implementation at FMPR, alongside their self-perceived EL readiness. Utilizing an exploratory, observational, monocentric design, this research assesses EMI’s potential to enhance equitable professional opportunities and doctoral career prospects locally and globally. By focusing on a public medical faculty serving a socioeconomically and educationally diverse student population, this study offers actionable insights applicable to comparable Global South contexts.

## Methods

### Study Design

This research is an exploratory, observational, monocentric cross-sectional study conducted at the FMPR during the 2023-2024 academic year. The study adhered to the Strengthening the Reporting of Observational Studies in Epidemiology (STROBE) guidelines. The primary objective was to explore the potential implementation of EMI at the FMPR. Specifically, the study aimed to analyze students’ perceptions and attitudes toward EMI, including its perceived benefits and disadvantages. In this context, we investigated students’ exposure to EL and their perceived EL proficiency in the four skills (listening, reading, speaking, and writing) in both general and academic EL to assess their readiness for the EMI curriculum. An exploratory design was chosen because there is no prior research on EMI implementation within a Moroccan public medical faculty. The observational aspect focused on collecting descriptive data from the sample, employing a quantitative approach using a questionnaire with Likert-type scale items and multiple-choice questions. Based on the objectives outlined above, the research questions were as follows:

- What is the perceived EL proficiency of students across the four EL skills, and more specifically in academic EL?
- What are the students’ perceptions and attitudes toward the potential implementation of EMI at the FMPR?
- What factors underpin students’ preference for EMI (EMI choice)?

### Participants and Setting

The target population consisted of medical students enrolled at the FMPR during the 2023-2024 academic year. A convenience sampling method was utilized, resulting in a sample of 102 undergraduate medical students from the second cycle of the medical curriculum (3rd, 4th, 5th, and 6th years of study). This sample size was considered appropriate for an exploratory study aimed at identifying preliminary trends. The inclusion criteria encompassed all enrolled students proficient in either FL or EL. The exclusion criteria were limited to students who declined to participate or who submitted incomplete questionnaires.

### Study Instrument

Data were collected using a custom-developed questionnaire divided into four sections. The first section gathered demographic data. The second section assessed students’ perceived proficiency in English, including their background, usage in general and academic contexts, and exposure across the four main language skills. The third section explored perceptions and attitudes toward EMI. The fourth section investigated students’ future EMI needs. The present study reported exclusively on data collected from the first three sections.

The questionnaire’s validity and reliability were established through rigorous, complementary methods. First, content and face validity were established through pilot testing among 15 participants from the target group, who assessed the items for clarity, relevance, and comprehensibility. Expert evaluation was subsequently conducted by six specialists, including three medicine and methodology professors and three experts in English studies and EMI. The questionnaire was iteratively refined based on their recommendations until final approval.

To assess the construct validity and underlying dimensional structure of the custom-developed questionnaire, an Exploratory Factor Analysis (EFA) was conducted using maximum likelihood extraction with varimax rotation. The suitability of the data for factor analysis was confirmed by a Kaiser-Meyer-Olkin (KMO) measure of sampling adequacy of 0.782 and a significant Bartlett’s test of sphericity (χ² (1035) = 3022, *P* < .001). The analysis confirmed the psychometric structure of the instrument, demonstrating proper item-scale alignment and adequate dimensionality (Table 1). Following this, the internal consistency reliability of the items in each scale was assessed using Cronbach’s alpha, utilizing a cut-off score of 0.50, which is considered acceptable for exploratory research [26,27]. Table 2 presents the alpha coefficients for each scale.

**Table 1.** EFA Psychometric Indicators.

| Measure | Value |
| --- | --- |
| **KMO** | 0.782 |
| **Bartlett's $\chi^2(df)$** | 3022 (1035) $p < .001$ |
| **Factor loadings** | >0.40 |

**Table 2 :**
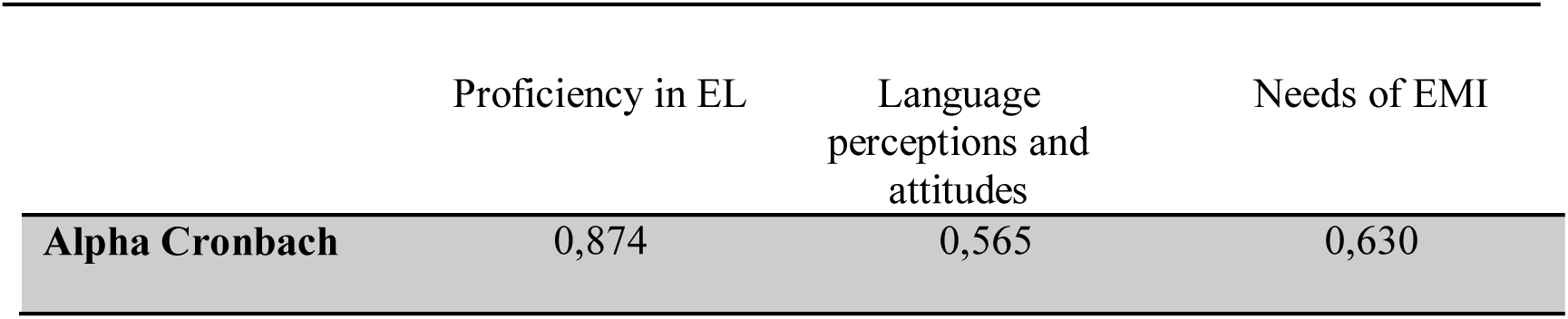
Scores Alpha Cronbach of questionnaire dimensions.

|  | Proficiency in EL | Language perceptions and attitudes | Needs of EMI |
| --- | --- | --- | --- |
| **Alpha Cronbach** | 0,874 | 0,565 | 0,630 |

### Data Collection Procedure

Data collection was conducted using a self-administered questionnaire. The questionnaire was edited in both FL and EL, allowing respondents to choose their preferred language version to maximize comprehension and response validity. A total of 130 questionnaires were distributed. Of these, 28 were incompletely answered and thus excluded, leaving 102 valid responses for the final analysis.

### Ethical Considerations

According to the guidelines of the Ethics Committee for Biomedical Research (CERB) at the FMPR, the committee’s mandate is to review biomedical research involving human subjects, such as clinical trials and studies involving medical interventions or the collection of biomedical patient data. Nonbiomedical research, including educational and social science surveys that do not involve clinical interventions, falls outside the scope of CERB review. Accordingly, this observational educational study did not require formal submission to or approval from the CERB. Nevertheless, all appropriate ethical precautions were strictly observed in accordance with the Declaration of Helsinki. All participants provided informed consent prior to participating. No identifying or sensitive personal information was collected; responses were submitted and gathered anonymously to ensure individual participants could not be traced.

### Statistical Analysis

Statistical analysis was performed using the open-source software Jamovi (version 2.6.26; The jamovi project) [28]. Prior to analysis, items were aggregated into dimension mean scores to measure the intended constructs: perceived proficiency in EL, and language perceptions and attitudes toward EMI. Descriptive statistics were calculated for all variables. Quantitative variables with non-Gaussian distributions were reported as medians and interquartile ranges, whereas qualitative variables were expressed as frequencies and percentages. Bivariate analyses were conducted using the chi-square test or Fisher exact test, as appropriate. Univariate and multivariate logistic regression models were employed to determine the effect of the explanatory variables on the dependent variable “EMI choice”. A *P* value < .05 was considered statistically significant. Associations were expressed as odds ratios (ORs) with 95% confidence intervals (CIs).

## Results

### Participant Characteristics

The sample comprised 102 participants, including 65 (63.7%) females and 37 (36.3%) males. The median age was 21 (IQR 21-23) years, with a range of 20 to 25 years. Regarding academic progression, 39 (38.2%) students were in their third year of study, 28 (27.5%) in their fourth year, 20 (19.6%) in their fifth year, and 14 (13.7%) in their sixth year. Demographic characteristics are summarized in Figures 1 and 2.

**Figure1.**
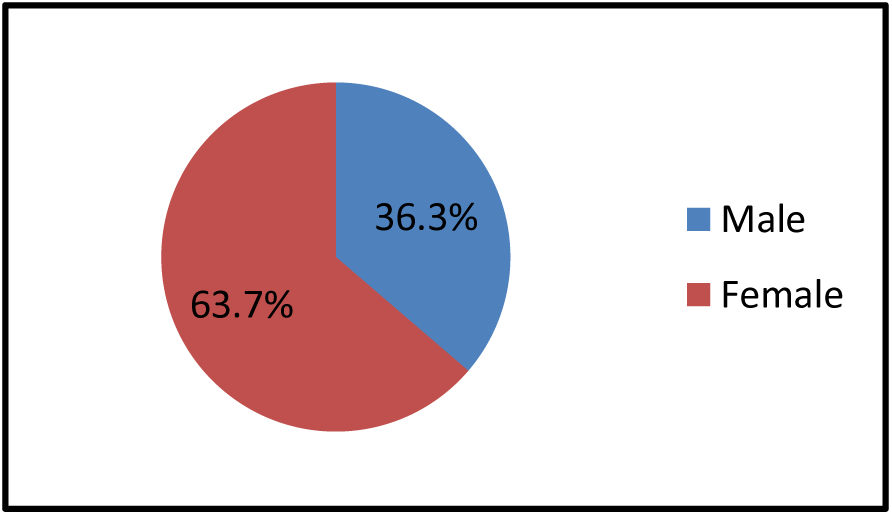
Distribution of students by gender

**Figure 2.**
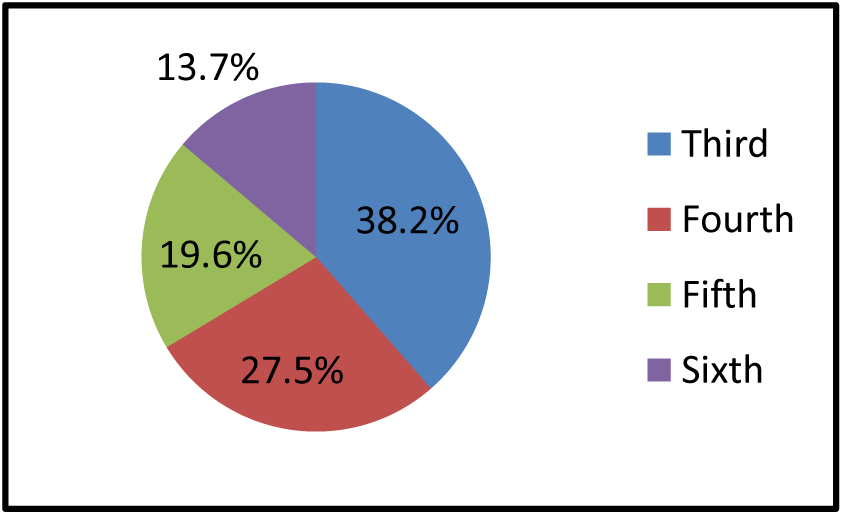
Distribution of students by year of study

The majority of participants (n=95, 93.1%) reported speaking EL in their daily lives. Over half (n=53, 52.0%) started learning EL in primary school, while 35 (34.3%) began in secondary school. However, only 29 (28.4%) students held an EL certification aligned with the Common European Framework of Reference for Languages (CEFR), specifically at B2 (n=10, 9.8%), C1 (n=10, 9.8%), and C2 (n=5, 4.9%) levels.

### Exposure to EL

Participants reported “moderate to frequent exposure” to EL (7 items; mean score 2.28/3, range reporting “often” 12.7%-87.3%; Table 3). All participants (102/102, 100%) declared frequently (“often”) practicing speaking and listening skills through habits such as interacting with friends, watching television, and listening to English music or podcasts. Conversely, exposure to reading and writing in EL was reported as “rare” (Table 3).

**Table 3.** Students with exposure to EL (n = 102)

|  | **Rarely**<br>**n (%)** | **Sometimes**<br>**n (%)** | **Often**<br>**n (%)** |
| --- | --- | --- | --- |
| **Interacting with friends** | 18 (17.6) | 32 (31.4) | 52 (51) |
| **Watching movies** | 5 (4.9) | 8 (7.8) | 89 (87.3) |
| **Listening to the radio/music** | 6 (5.9) | 8 (7.8) | 88 (86.3) |
| **Listening to podcasts** | 10 (9.8) | 25 (24.5) | 67 (65.7) |
| **Reading** | 13 (12.7) | 40 (39.2) | 49 (48) |
| **Free writing** | 53 (52) | 32 (31.4) | 17 (16.7) |
| **Private English classes** | 69 (67.6) | 20 (19.6) | 13 (12.7) |

### Perceived EL Proficiency

Regarding general EL competencies, self-reported proficiency was largely positive (4 items; mean score 2.58/3, range rating “good” 60.8%-75.5%). The majority of participants evaluated their abilities as “good” across routine tasks, including listening, reading, writing on social media, and speaking during everyday interactions.

In contrast, perceptions of academic EL proficiency were notably lower (7 items; mean score 2.05/3, range rating “average” to “good” 19.6%-53.9%; Table 4). Participants rated their receptive skills (listening and reading) as “good”, whereas their productive skills (speaking and writing) were considered “average”. Specifically, the majority of students considered their ability to write a scientific research article as “basic”.

**Table 4.** Perceived students’ skills in General and Academic EL (n = 102)

|  | **Basic**<br>**n(%)** | **Average**<br>**n (%)** | **Good**<br>**n (%)** |
| --- | --- | --- | --- |
| ***General EL*** |  |  |  |
| **Speaking in regular everyday interactions** | 10 (9.8) | 30 (29.4) | 62 (60.8) |
| **Listening in regular daily interactions** | 7 (6.9) | 18 (17.6) | 77 (75.5) |
| **Reading about general topics of interest to me (Newspapers, magazines, books)** | 9 (8.8) | 26 (25.5) | 67 (65.7) |
| **Communicate in writing on social media platforms** | 7 (6.9) | 26 (25.5) | 69 (67.6) |
| ***Academic EL*** |  |  |  |
| **Discuss technical ideas in the field of medicine** | 29 (28.4) | 46 (45.1) | 27 (26.5) |
| **Prepare and deliver a presentation about a medical issue** | 31 (30.4) | 47 (46.1) | 24 (23.5) |
| **Understand videos about medical issues** | 11 (10.8) | 36 (35.3) | 55 (53.9) |
| **Read medicine textbooks written in EL** | 17 (16.7) | 36 (35.3) | 49 (48) |
| **Write class assignments in EL about topics in medicine** | 25 (24.5) | 50 (49) | 27 (26.5) |
| **Communicating in EL with my course instructors** | 30 (29.4) | 45 (44.1) | 27 (26.5) |
| **Write a scientific article for publication in a medical journal** | 44 (43.1) | 38 (37.3) | 20 (19.6) |

A minority of respondents expressed apprehension regarding specific academic tasks, rating their skills as “basic” for understanding videos on medical issues (n=11, 10.8%), comprehending English-medium medical textbooks (n=17, 16.7%), writing academic assignments in EL (n=25, 24.5%), participating in class discussions (n=29, 28.4%), and preparing and delivering presentations (n=31, 30.4%).

### Perceptions and Attitudes toward EL and EMI

Participants demonstrated favorable perceptions and attitudes toward EL and EMI implementation in medical education (9 items; mean score 2.83/3, range of “agree” 64.7%-98.0%; Table 5). 98 (96.1%) respondents agreed that EL is useful in the medical field, and 90 (88.2%) believed that EL should be the language of higher education. However, 74 (72.5%) students expressed a preference to maintain a bilingual medical curriculum, utilizing both FL and EL as media of instruction at the faculty.

**Table 5.** Students ‘perceptions and attitudes (n = 102)

|  | **Agree**<br>**n (%)** | **Neutral**<br>**n (%)** | **Disagree**<br>**n (%)** |
| --- | --- | --- | --- |
| **I like to learn EL** | 91 (89.2) | 7 (6.9) | 4 (3.9) |
| **I like to use EL** | 85 (83.3) | 15 (14.7) | 2 (2) |
| **EL is very useful for work in Morocco** | 66 (64.7) | 35 (34.3) | 1 (1) |
| **EL is useful for medical scientific field** | 98 (96.1) | 4 (3.9) | - |
| **EL should be the language of higher education** | 90 (88.2) | 11 (10.8) | 1 (1) |
| **We should have more courses using EMI in higher education** | 89 (87.3) | 13 (12.7) | - |
| **If an EMI program is offered in my faculty, I will definitely go for it.** | 84 (82.4) | 18 (17.6) | - |
| **Multilingualism offers me advantages** | 100 (98) | 2 (2) | - |
| **We should keep both FL and EL as medium of instruction in higher education** | 74 (72.5) | 25 (24.5) | 3 (2.9) |

### Perceived Benefits and Disadvantages of EMI

Participants recognized significant advantages to EMI implementation (4 items; mean score 2.48/3, range of “yes” 65.7%-83.2%; Table 6). Specifically, respondents indicated that EMI would facilitate international mobility (n=85, 83.3%) provide better professional opportunities (n=82, 80.4%), enhance the learning of medical content (n=71, 69.6%), and improve their EL skills (n=67, 65.7%).

**Table 6.**
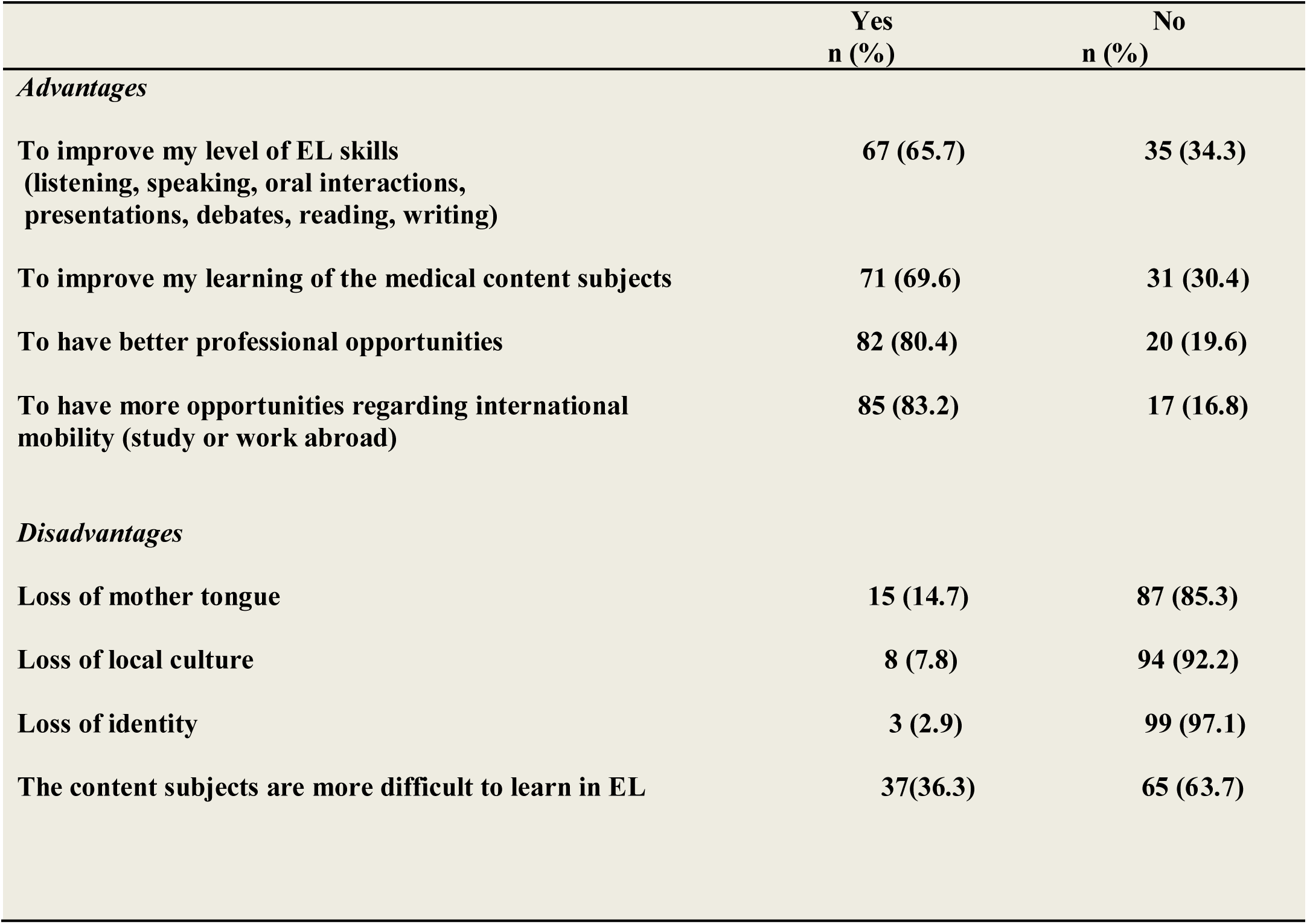
Perceived benefits and disadvantages of EMI according to students (n = 102)

Conversely, perceived disadvantages received lower rates of agreement (4 items; mean score 1.30/3, range of “yes” 2.9%-36.3%; Table 6). Reported concerns included the potential difficulty of learning medical subjects in EL (n=37, 36.3%), the erosion of the mother tongue (n=15, 14.7%), the loss of local culture (n=8, 7.8%), and the loss of personal identity (n=3, 2.9%).

A bivariate analysis was conducted to examine the association between the explanatory variables and the dependent variable representing EMI choice (“If an EMI course is offered in my faculty, I will definitely go for it”). Statistically significant associations were observed between EMI choice and age (*P* = .035), course year (*P* < .001), the variable “I like learning EL” (*P* = .01), and the variable “EL should be the language of higher education” (*P* < .001) (Table 7). The proportion of EMI choice increased with academic progression, rising from 53.8% in the third year to 96.4% in the fourth year. High percentages of EMI preference w also were also observed among students who agreed with “I like learning EL” (75.8%) and EL should be the language of higher education” (80.5%). Gender and EL certification did not reach statistical significance.

**Table 7.**
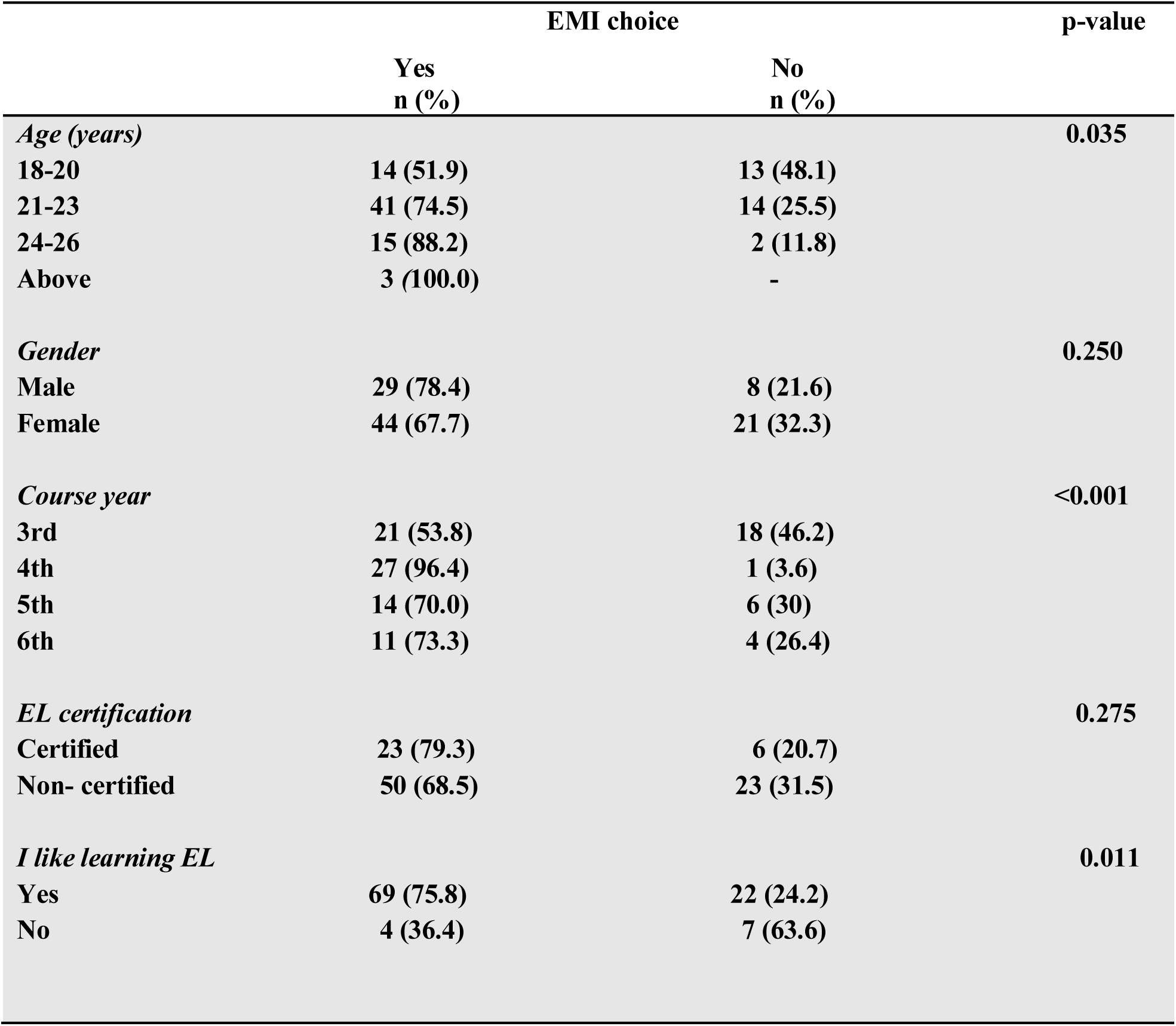
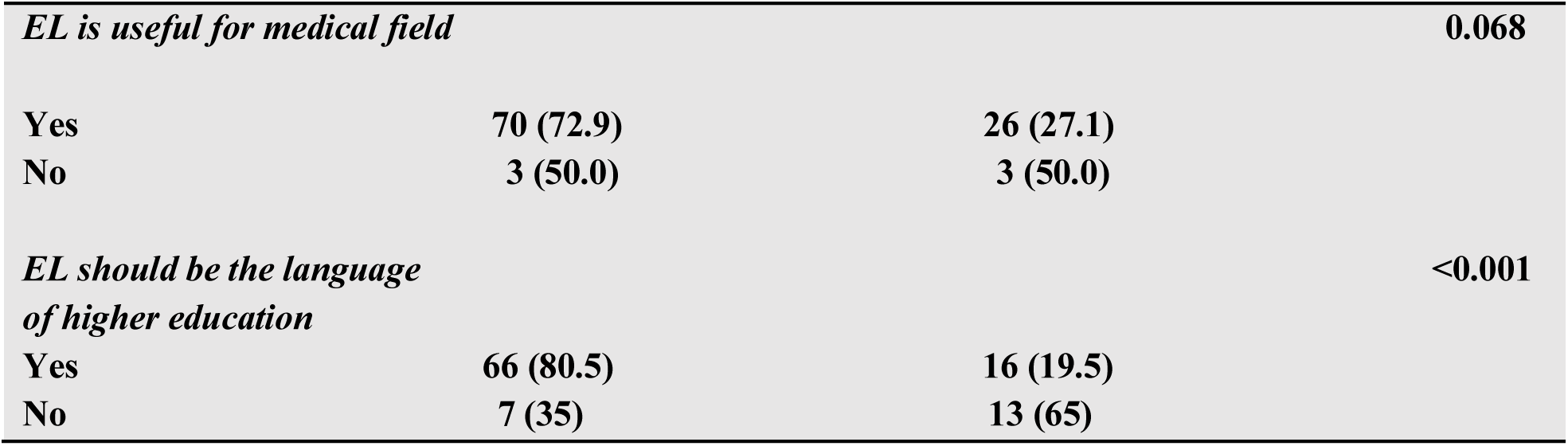
Bivariate analysis between EMI choice and the explanatory variables (n = 102)

For the variable “EL is useful for the medical field”, 96.1% of respondents agreed with the statement, and 72.9% of those who agreed chose EMI. The primary *P* value for this variable did not reach statistical significance, a mathematical outcome attributed to the strong homogeneity of responses and subsequent reduced variance. Given the resulting low expected cell counts, the Fisher exact test was utilized. Calculation of the Cramer V coefficient revealed a moderate mathematical association (Cramer V = 0.207).

To further evaluate predictors of EMI choice, univariate and multivariate logistic regression analyses were performed (Table 8). The univariate regression identified three statistically significant predictors: course year 4 vs 3 (OR 23.14, 95% CI 2.85-187.60; *P* = .003), “I like learning EL” (OR 5.48, 95% CI 1.46-20.52; *P* = .01), and “EL should be the language of higher education” (OR 7.66, 95% CI 2.63-22.31; *P* < .001).

**Table 8.** Associated factors with EMI choice: Univariate and multivariate logistic regression models.

| Associated factors | Univariate model |  |  | Multivariate model |  |  |
| --- | --- | --- | --- | --- | --- | --- |
|  | OR | CI 95% | p-value | OR | CI 95% | p-value |
| **Gender** | **0.578** | **0.22 - 1.48** | **0.253** | **0.31** | **0.08 - 1.10** | **0.071** |
| **Course year** |  |  |  |  |  |  |
| 4 – 3 | 23.14 | 2.85 - 187.6 | 0.003 | 8.83 | 2.05 - 172.69 | 0.009 |
| 5 – 3 | 2.00 | 0.63 - 6.28 | 0.235 | 1.81 | 0.50 - 6.49 | 0.36 |
| 6 – 3 | 2.36 | 0.63 - 8.70 | 0.198 | 2.13 | 0.44 - 10.15 | 0.34 |
| **EL certification** | **1.76** | **0.63 - 4.92** | **0.278** | **1.72** | **0.49 - 6.02** | **0.395** |
| **I like learning EL** | **5.48** | **1.46 - 20.52** | **0.011** | **6.90** | **1.31 - 36.16** | **0.022** |
| **EL is useful for medical field** | **2.69** | **0.51 - 14.19** | **0.243** | **1.13** | **0.12 - 10.51** | **0.911** |
| **EL should be the language of higher education** | **7.66** | **2.63 - 22.31** | **<0.001** | **6.38** | **1.87 - 21.72** | **0.003** |

Subsequently, a backward stepwise multivariate regression was conducted to retain the most relevant predictors based on their statistical significance. Two additional variables, gender and “EL is useful for the medical field” were forced into the model based on their contextual and theoretical relevance. In the final multivariate model, the three primary variables maintained a statistically significant positive influence on EMI choice: course year 4 vs 3 (OR 8.83, 95% CI 2.05-172.69; *P* = .009), “I like learning EL” (OR 6.90, 95% CI 1.31-36.16; *P* = .02), and “EL should be the language of higher education” (OR 6.38, 95% CI 1.87-21.72; *P* = .003). The two variables included based on theoretical relevance did not reach statistical significance in the final model.

## Discussion

This study revealed generally positive attitudes among medical students at the FMPR toward the implementation of EMI. Participants recognized the profound utility of EL for the medical field, valuing its role in fostering international mobility, expanding professional opportunities, and enhancing the learning of medical content. Crucially, this strong support within FMPR’s socioeconomically diverse public population institution challenges the critique that EMI is solely a private privilege, pointing instead to the potential of public EMI programs to promote educational democratization.

The favorable perspective of the respondents aligns with established arguments supporting EMI in medical education. Because international medical communication and literature predominantly occurs in EL, EMI enables students to access the latest clinical studies, stay updated with evidence-based practices, and collaborate with the global medical community, which is pivotal for professional growth [5]. The data demonstrated a significant developmental trend: as students advanced in their medical training, their preference for an EMI curriculum increased substantially, reflecting the pressing need to access global medical literature for their graduation theses and postgraduate specialization. Furthermore, intrinsic motivation to learn EL and a preference for EL as the primary language of higher education were strong predictors of EMI acceptance. Such positive dispositions facilitate the acquisition of complex medical terminology, which is indispensable for international scientific exchange. This aligns with Nguyen who pointed out that an EMI curriculum facilitates the acquisition of medical terminology, an indispensable prerequisite for international scientific exchange [29]. Similarly, studies from Saudi Arabia reported comparable favorable attitudes toward EMI in scientific disciplines, with medical students expressing a strong willingness to study using EL [30,31].

A critical insight from this study is the particularity of EMI programs in public medical faculties, where students originate from diverse social backgrounds and may not have had equal prior educational opportunities. Highlighting this complexity, Roshini noted that linguistic background plays a key role in academic performance [32]. Unlike private institutions where EMI often serves privileged students, the socioeconomic diversity at the FMPR reveals a broader equity potential. A vast majority of students, including those from less affluent backgrounds, acknowledged the career benefits of an EL medium medical training. These results suggest that integrating EMI in public institutions could mitigate existing institutional disparities, fostering thus educational equity, and democratizing access to global opportunities, such as WFME and ECFMG accreditations, historically dominated by graduates of elite private institutions [24,25].

The positive attitudinal profiles of Moroccan medical students coincide with a pivotal historical juncture. Following the Brexit, an agreement between Morocco and the United Kingdom, signed on 26 October 2019, entered into force on 1 January 2021. In the educational field, one of the major Moroccan goals of this British partnership is to enhance its attractiveness for researchers and PhD students in the UK [33]. To align with this goal, several Moroccan private universities, such as Mohammed VI Polytechnic University in Benguerir (UM6P) and University Mohammed VI of Health Sciences (UM6SS) introduced EMI medical programs [34,35]. However, restricting EMI to the private universities may exacerbate social inequalities. To address this, Morocco and UK signed in a 2023 Memorandum of Understanding (MoU) aiming to increase EL integration in Moroccan public universities [37]. While this public-private educational disparity mirrors global trends identified in British Council surveys, this study underscored the specific sectoral challenges within public medical education. Despite these challenges, the FMPR data revealed a remarkable student receptiveness toward transitioning to EMI [6].

In the context of global EMI trends, Moroccan students’ positive attitudes align with those of their regional peers in the MENA region, such as Algeria and Libya [20,37], as well as broader African contexts [38]. This enthusiasm is equally comparable to trends observed in high-income countries like Japan [3]. Nevertheless, proficiency gaps remain a threat to achieving equitable outcomes [15]. These insights could inform EMI policy development in comparable medical public institutions, advocating hybrid EMI integration models to address pedagogical challenges while capitalizing on students’ favorable attitudes to enhance global competitiveness.

The receptiveness to EMI is strongly supported by a young population that is highly favorable toward EL [39]. For Moroccan youth, EL is the language of digital media and popular culture (Social networks, movies and music), which facilitates the robust development of Basic Interpersonal Communication Skills (BICS) [40]. However, this conversational fluency does not automatically translate to the Cognitive Academic Language Proficiency (CALP) required for medical EMI. Students’ daily engagement with EL correlates with their intrinsic motivation. A similar enthusiasm was documented by Belhiah in a multicenter study where students expressed a passion for EL as a subject worth studying [41].

More broadly, this enthusiasm aligns with an ongoing reorientation of language-in-education policies across North Africa [10]. Despite a shared colonial legacy that historically established FL as the dominant language of instruction, there is a pragmatic transition toward EL as the global lingua franca of medicine [10]. At the institutional level, however, practical concerns persist among some students regarding the dual cognitive load of simultaneously mastering complex medical content and improving their proficiency in EL as the new medium of instruction, underscoring the need for structured pedagogical support within EMI programs [42]. Conversely, cultural concerns regarding the loss of mother tongue or local identity were notably minimal. These relatively minimal cultural concerns may reflect the fact that the medium of higher education is already a non-native language, FL, which has historically not been perceived as having a detrimental impact on Moroccan students’ cultural identity. Within this context, EL tends to be considered primarily as a professional resource. Unlike FL, which carries historical and colonial connotations and has traditionally functioned as a marker of elite status, EL is widely perceived in Morocco as a relatively neutral gateway to international mobility and medical technologies [43]. Consequently, students’ multilingualism appears to foster an additive approach to language acquisition, whereby EL is embraced mainly for its instrumental value in facilitating global medical integration.

### Limitations and future research directions

This study has limitations that should be acknowledged. First, the findings are based on self-reported perceptions, which may not fully reflect students’ actual EL proficiency or academic performance under EMI conditions. Second, the cross-sectional design captures attitudes at a single point in time and does not allow examination of how perceptions evolve across the medical training continuum. Third, the monocenter sample, while providing detailed insights into the FMPR, limits the generalizability of the results to other Moroccan medical schools, regions, or disciplines.

The absence of qualitative data represents an additional important limitation. Although the questionnaire included an open-ended question, no participants responded, leaving the subjective experiences behind the quantitative patterns unexplored. Future research should therefore incorporate qualitative approaches, such as semi-structured interviews or focus groups, to investigate students’ lived experiences, perceived challenges, and views on the impact of EMI on their learning and performance after implementation. Such in-depth insights could identify unanticipated issues and inform targeted curriculum design and support strategies.

Moreover, the observed association between EMI choice and the perception that EL is useful for the medical field, although statistically meaningful (Cramer’s V = 0.207), is modest in strength and suggests further investigation. Mixed-methods or longitudinal designs could help clarify how and why this relationship emerges and which contextual or individual factors may mediate or moderate it.

Despite these limitations, the study provides valuable insights into EMI-related perceptions within a specific regional context. By elucidating trends among medical students in a public Moroccan faculty, it contributes to the growing evidence base on EMI in medical education and may inform research, policy, and practice in comparable educational and cultural settings where EL is increasingly integrated into medical education worldwide.

### Conclusion

The findings of this study suggest that integrating EMI into public medical faculties in Morocco is perceived more as an opportunity than as a constraint. Most participating second-cycle students at the FMPR expressed positive attitudes toward EMI and generally favorable self-perceptions of their EL proficiency, particularly regarding general and academic listening and reading skills. Support for EL and EMI was stronger among students in higher academic years, accompanied by heightened awareness of the role of EL in international professional development and scientific engagement in medicine. EL was predominantly framed as a pragmatic professional resource and global lingua franca, rather than as a language associated with colonial connotations, which may help explain its broad acceptance among students from diverse social backgrounds.

Within this context, EMI holds potential to support a more equitable distribution of educational and professional opportunities for medical students across different types of institutions. At the same time, the observed discrepancy between strong enthusiasm for EMI and self-reported weaker EL proficiency in a subset of students underscores the need for implementation strategies that are responsive to student needs. Institutional strategies should therefore include structured linguistic and pedagogical support to reduce additional cognitive and academic burdens and to prevent the widening of existing inequities. Overall, EMI in Moroccan public medical education appears to offer a credible pathway to greater equity and global competitiveness, provided that its integration is gradual, adequately resourced, and responsive to local needs and constraints.

## Data Availability

We have included a data availability statement in the revised manuscript as requested: The datasets generated and analyzed during the study are available from the corresponding author. Due to ethical considerations and participant confidentiality, the data are not publicly available.

## Conflicts of interest

None.

## List of abbreviations

AL: Arabic language
BICS: Basic Interpersonal Communication Skills
CALP: Cognitive Academic Language Proficiency
CEFR: Common European Framework of Reference for Languages
CERB: Ethics Committee for Biomedical Research
ECFMG: Educational Commission for Foreign Medical Graduates
EFA: Exploratory Factor Analysis
EL: English language
EMI: English as medium of instruction
FL: French language
FMPR: Faculty of Medicine and Pharmacy of Rabat
MoU: Memorandum of Understanding
STROBE: Strengthening the Reporting of Observational Studies in Epidemiology
UK: United Kingdom
UM6P: Mohammed VI Polytechnic University in Benguerir
UM6SS: University Mohammed VI of Health Sciences
USMLE: United States Medical Licensing Examination
WFME: World Federation for Medical Education

## Appendix : Questionnaire

Dear participant,

This questionnaire is part of a research project on the use of the English language (EL) as a means of instruction in your faculty (EMI: English as a medium of instruction). Some questions are in the form of Likert scales while others have multiple choices, allowing you to check the answers that seem most appropriate to you. Your contribution is invaluable to the completion of this research project and is highly appreciated. Your anonymous answers will be held in strict confidentiality and will be used only for the purposes of this study. The results will be reported in aggregate form only, and cannot be identified individually. Thank you for your collaboration.

The researcher

## I. DEMOGRAPHIC DATA

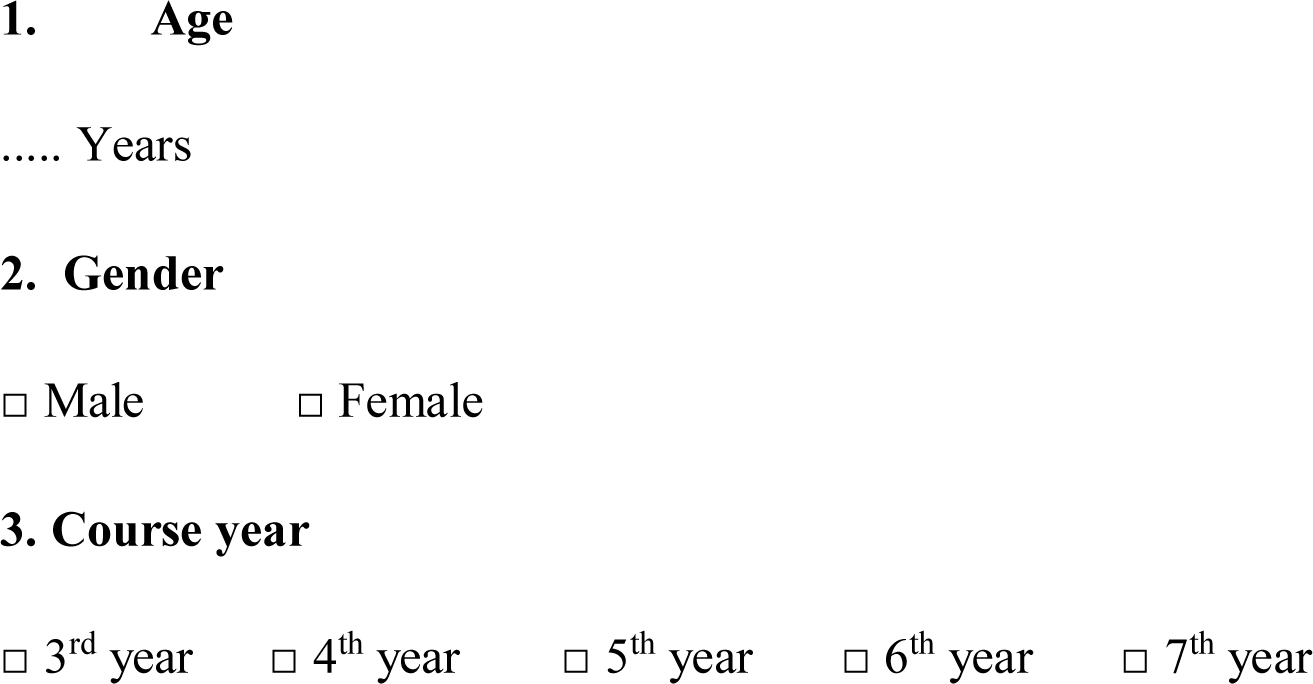

## II. PROFICIENCY IN ENGLISH LANGUAGE

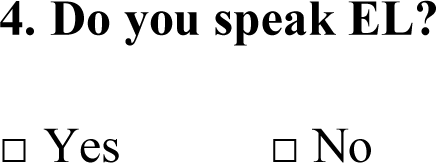
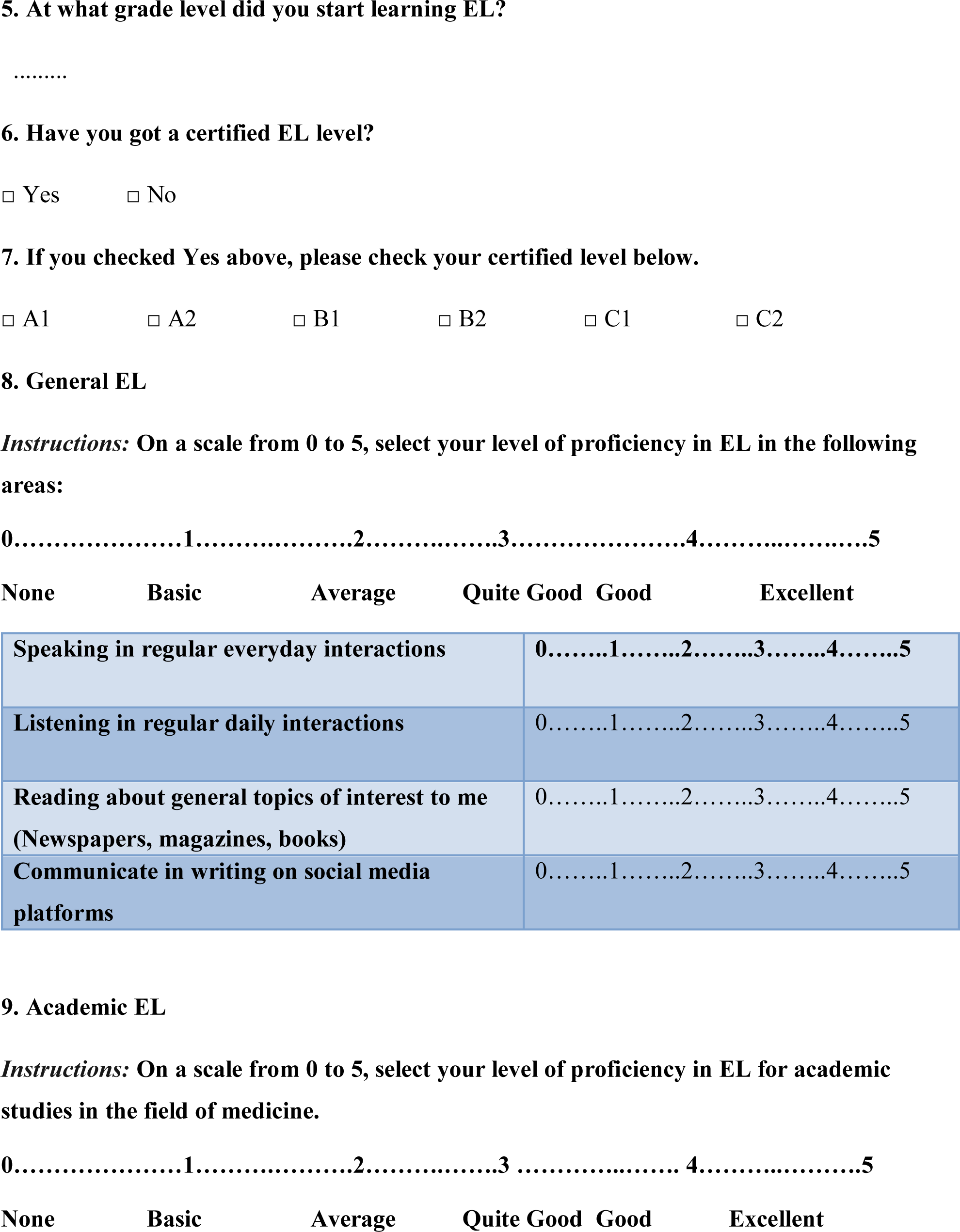
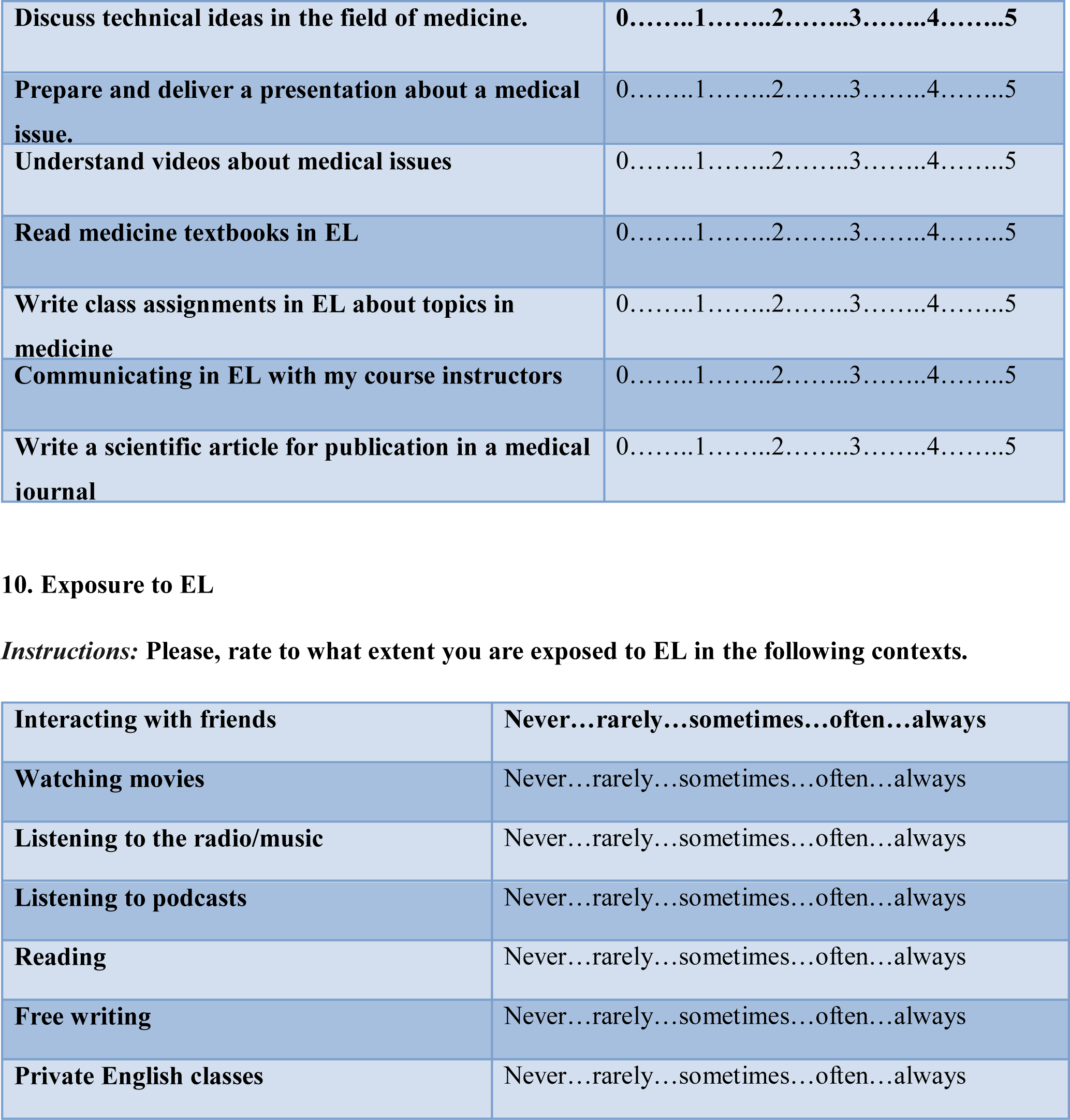
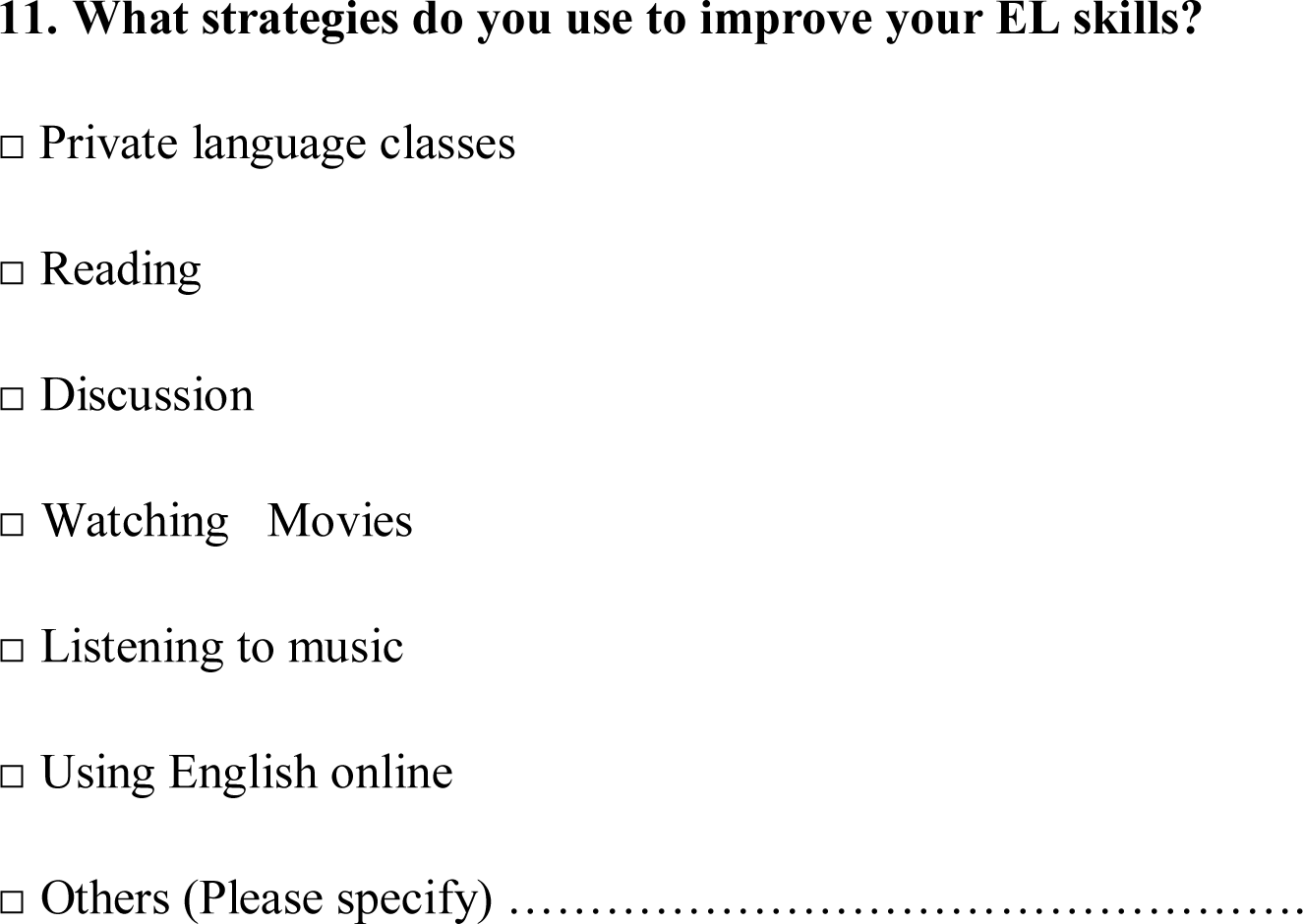

## III. LANGUAGE PERCEPTIONS AND ATTITUDES

**Strongly Disagree Disagree Neutral Agree Strongly Agree**
|  |  |
| --- | --- |
| **I like learning EL** | **1.....2.....3.....4.....5** |
| **I like to use EL** | **1.....2.....3.....4.....5** |
| **EL is very useful for work in Morocco** | **1.....2.....3.....4.....5** |
| **EL is useful for medical scientific field** | **1.....2.....3.....4.....5** |
| **EL should be the language of higher education** | **1.....2.....3.....4.....5** |
| **We should have more courses using EMI in higher education** | **1.....2.....3.....4.....5** |
| **If an EL program is offered in my faculty,**<br>**I will definitely go for it.** | 1.....2.....3.....4.....5 |
| **Multilingualism offers me advantages** | 1.....2.....3.....4.....5 |
| **We should keep both French language**<br>**and EL as mediums of instruction in**<br>**higher education** | 1.....2.....3.....4.....5 |

## IV. NEEDS OF EMI

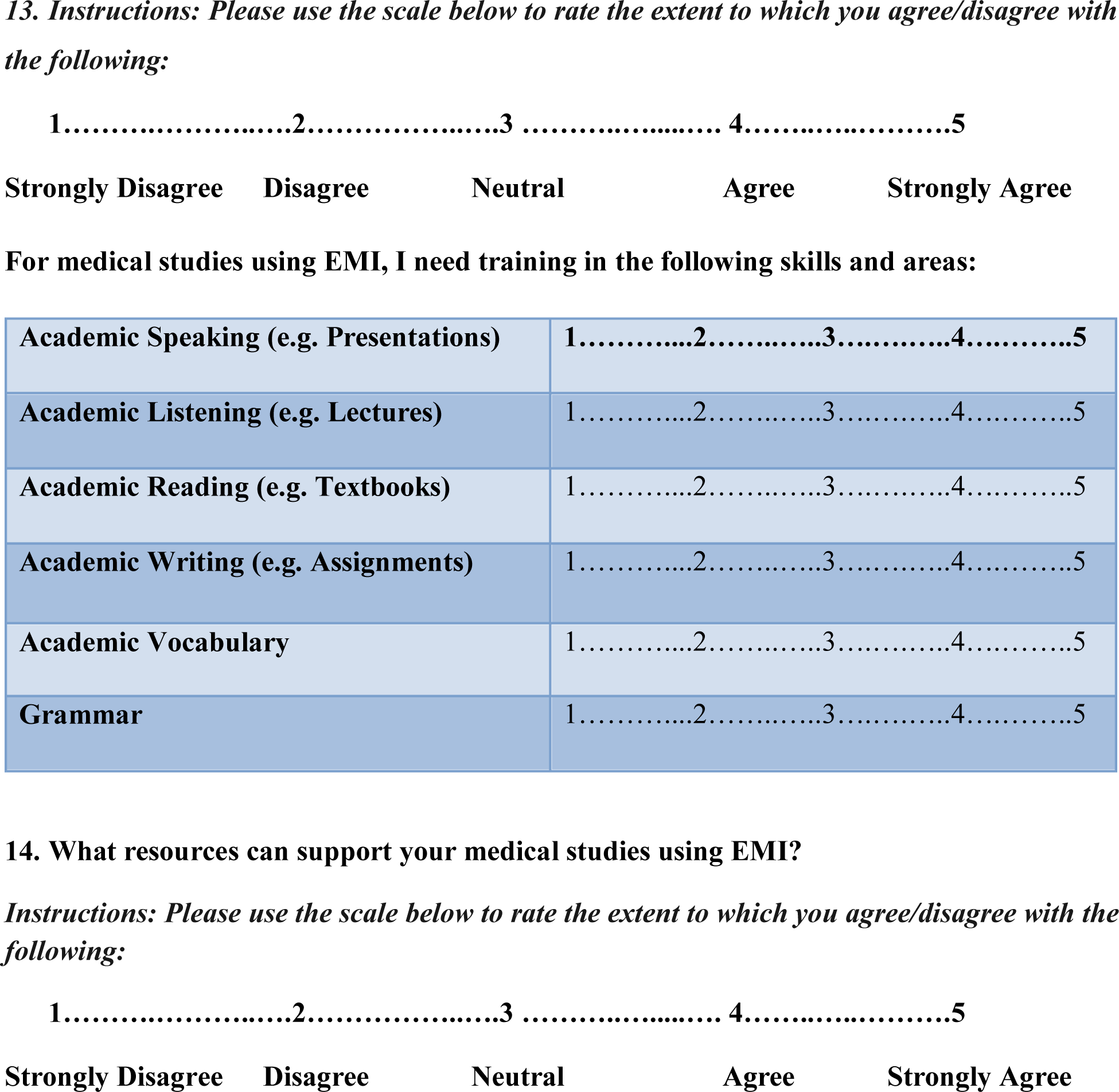
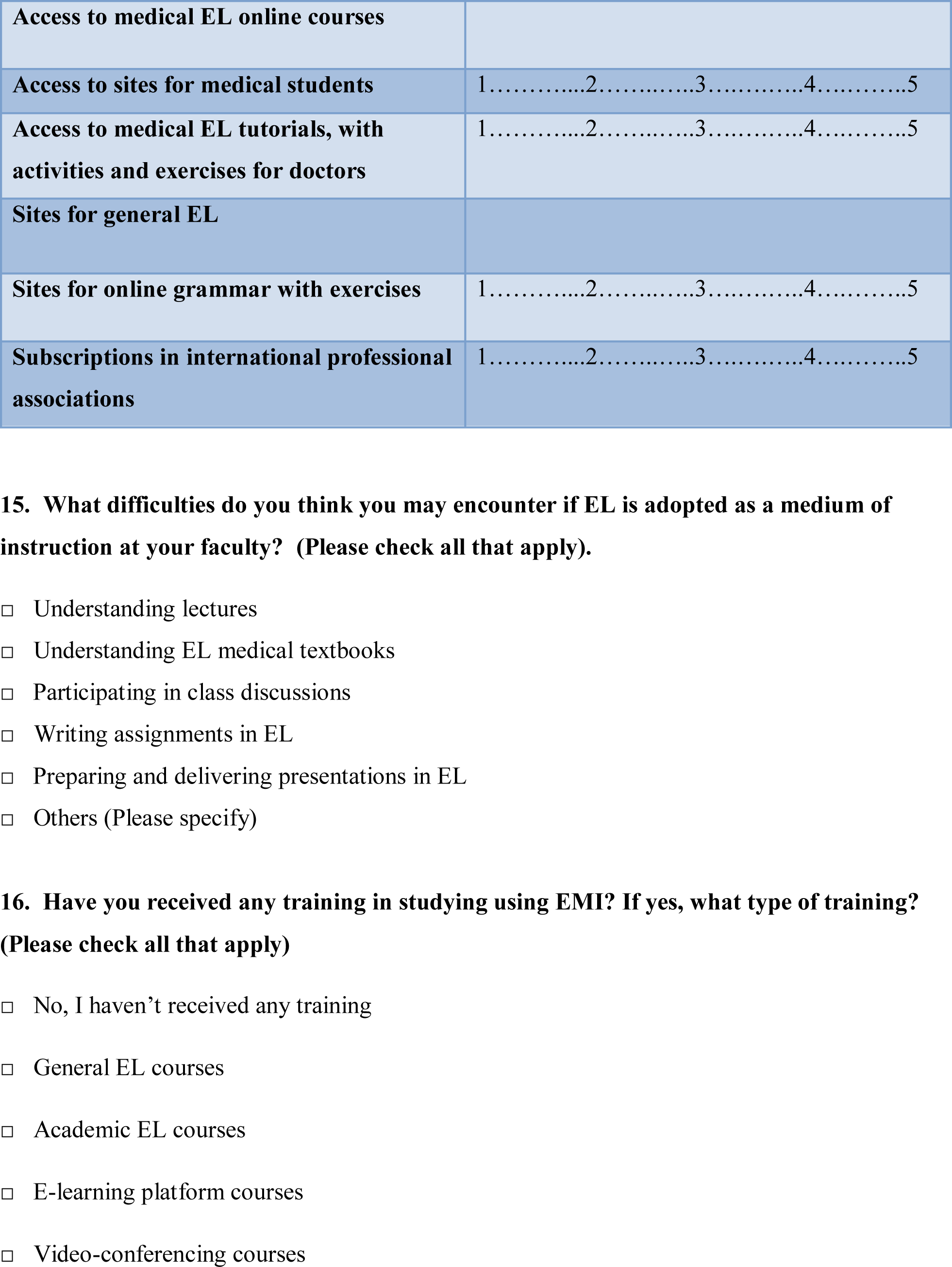
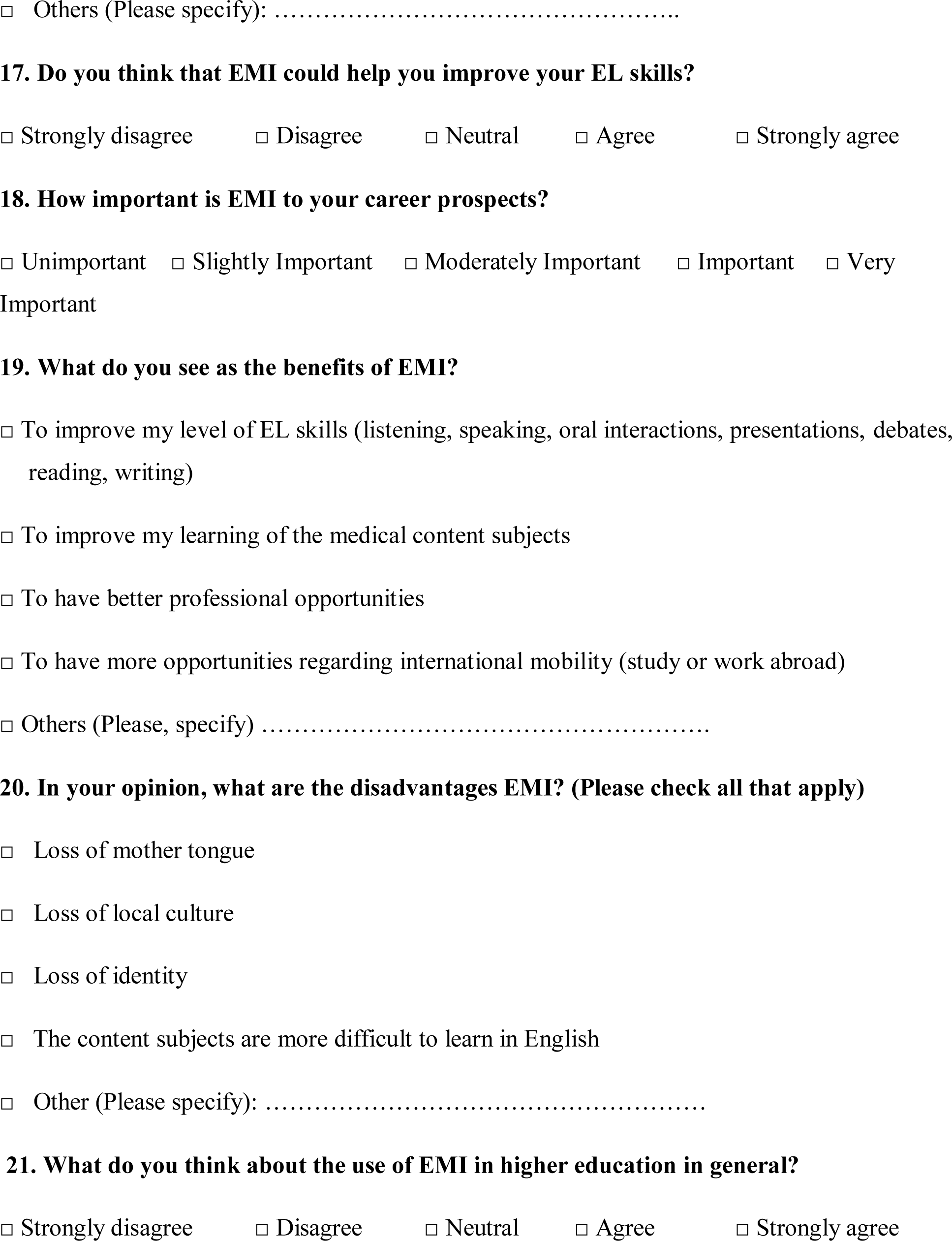
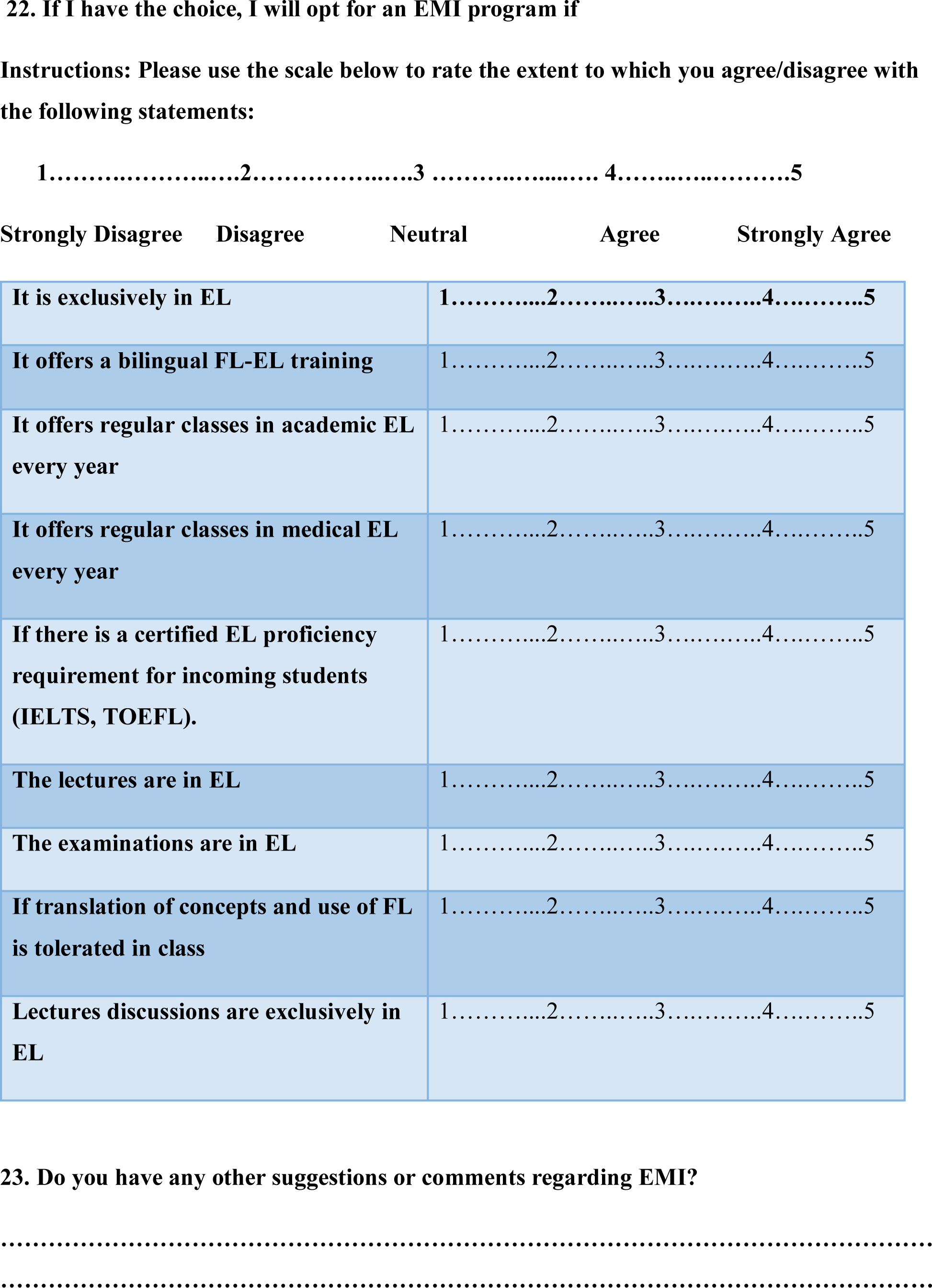

## Notes

### Competing Interest Statement

The authors have declared no competing interest.

### Funding Statement

This study did not receive any funding.

### Author Declarations

According to section 2 of the guidelines of the Committee of Ethics (CERB) at the Faculty of Medicine and Pharmacy of Rabat, the committee reviews only biomedical research projects involving human subjects, such as clinical trials, biomedical experiments, or studies directly related to human health. Preclinical research (in vitro or animal studies) and non-biomedical research, including studies related to social sciences or education, without medical interventions or biomedical data collection, are generally outside the scope of the CERBs review. Therefore, according to CERB policies and Moroccan regulations, this type of study does not require formal submission to or approval from the CERB. Nevertheless, although formal approval from the CERB was not specifically required for this study, we took all necessary ethical precautions, such as obtaining voluntary informed consent from all participants and ensuring the confidentiality and anonymity of the data collected. We confirm that the study adhered to the core ethical principles of the Declaration of Helsinki. Although our research involved human participants, it focused on non-interventional aspects related to perceptions and attitudes regarding English as a Medium of Instruction, and was conducted with respect for participant anonymity, informed consent, and minimizing risk. Furthermore, in line with applicable local regulations and CERBs guidelines, our research was guided by the ethical oversight of qualified personnel familiar with ethical research standards. We included in our study a statement regarding informed consent as part of our commitment to uphold ethical research standards, even though obtaining formal consent was not strictly required for this non-interventional study focused on language attitudes and perceptions. All participants voluntarily agreed to take part after being fully informed about the nature and purpose of the study. Participant confidentiality and anonymity were rigorously maintained throughout.

### Summary of Updates

Incorporated recent literature on EMI in Moroccan medical education ; minor revisions for clarity

